# Non-invasively measured brain activity and radiological progression in diffuse glioma

**DOI:** 10.1101/2021.01.31.21250524

**Authors:** T. Numan, S.D. Kulik, B. Moraal, J.C. Reijneveld, CJ. Stam, P.C. de Witt Hamer, J. Derks, A.M.E. Bruynzeel, M.E. van Linde, P. Wesseling, M.C.M. Kouwenhoven, M. Klein, T. Würdinger, F. Barkhof, J.J.G. Geurts, A. Hillebrand, L. Douw

## Abstract

Non-invasively measured brain activity is related to progression-free survival in glioma patients, suggesting its potential as a marker of glioma progression. We therefore assessed the relationship between brain activity and increasing tumor volumes on routine clinical magnetic resonance imaging (MRI) in glioma patients. Postoperative magnetoencephalography (MEG) was recorded in 45 diffuse glioma patients. Brain activity was estimated using three measures (absolute broadband power, offset and slope) calculated at three spatial levels: global average, averaged across the peritumoral areas, and averaged across the homologues of these peritumoral areas in the contralateral hemisphere. Tumors were segmented on MRI. Changes in tumor volume between the two scans surrounding the MEG were calculated and correlated with brain activity. Brain activity was compared between patient groups classified into having increasing or stable tumor volume. Results show that brain activity was significantly increased in the tumor hemisphere in general, and in peritumoral regions specifically. However, none of the measures and spatial levels of brain activity correlated with changes in tumor volume, nor did they differ between patients with increasing versus stable tumor volumes. Longitudinal studies in more homogeneous subgroups of glioma patients are necessary to further explore the clinical potential of non-invasively measured brain activity.

## Introduction

Monitoring treatment response and disease progression in diffuse glioma patients is challenging. The current standard for monitoring tumor growth is magnetic resonance imaging (MRI), namely T1-weighted scans before and after contrast injection, a T2-weighted scan and a fluid-attenuated inversion recovery (FLAIR) sequence^1^. However, determining growth can be challenging^2^. Moreover, other patient factors such as cognitive deterioration may precede radiological progression^3,4^, underlining the relevance of exploring alternative markers of growth.

There is a causal relationship between increased peritumoral neuronal activity (i.e. spiking rate) and accelerated glioma growth^5,6^. Moreover, glutamate-dependent ‘neurogliomal’ synapses are formed between healthy neurons and nearby glioma cells, resulting in interconnected glioma and neuronal networks^7,8^. Based on these studies, it seems that both higher spiking rates and greater glutamate-related excitation cause increased proliferation and invasion of glioma, at least in animal models. Translating these findings to patients, glioma-infiltrated brain regions showed greater neuronal activity measured with intraoperative electrocorticography as compared to healthy appearing regions in three glioblastoma (GBM) patients^8^, further supporting this association between brain and tumor activities.

In recent translational studies, magnetoencephalography (MEG) was established as a non-invasively measured proxy of neuronal activity^9^. MEG records the magnetic fields induced by (mainly) postsynaptic neuronal currents with high temporal and varying spatial resolution^10,11^. Each sensor’s time-series reflects brain activity across different frequencies. The power spectrum can be determined to reduce these multidimensional data to a single indication of brain activity. This spectrum reflects the squared amplitude of all frequencies (figure 1), and the broadband power is the sum of squared amplitudes across all frequencies. We previously found that higher global broadband power at diagnosis and after tumor resection relates to shorter progression-free survival (PFS^12,13^), also after taking known predictors into account, and may thus be an early marker of tumor progression. As a next step in the investigation of the clinical relevance of non-invasively measured brain activity, we aimed to relate brain activity to ongoing radiological glioma growth.

**Figure 1.**
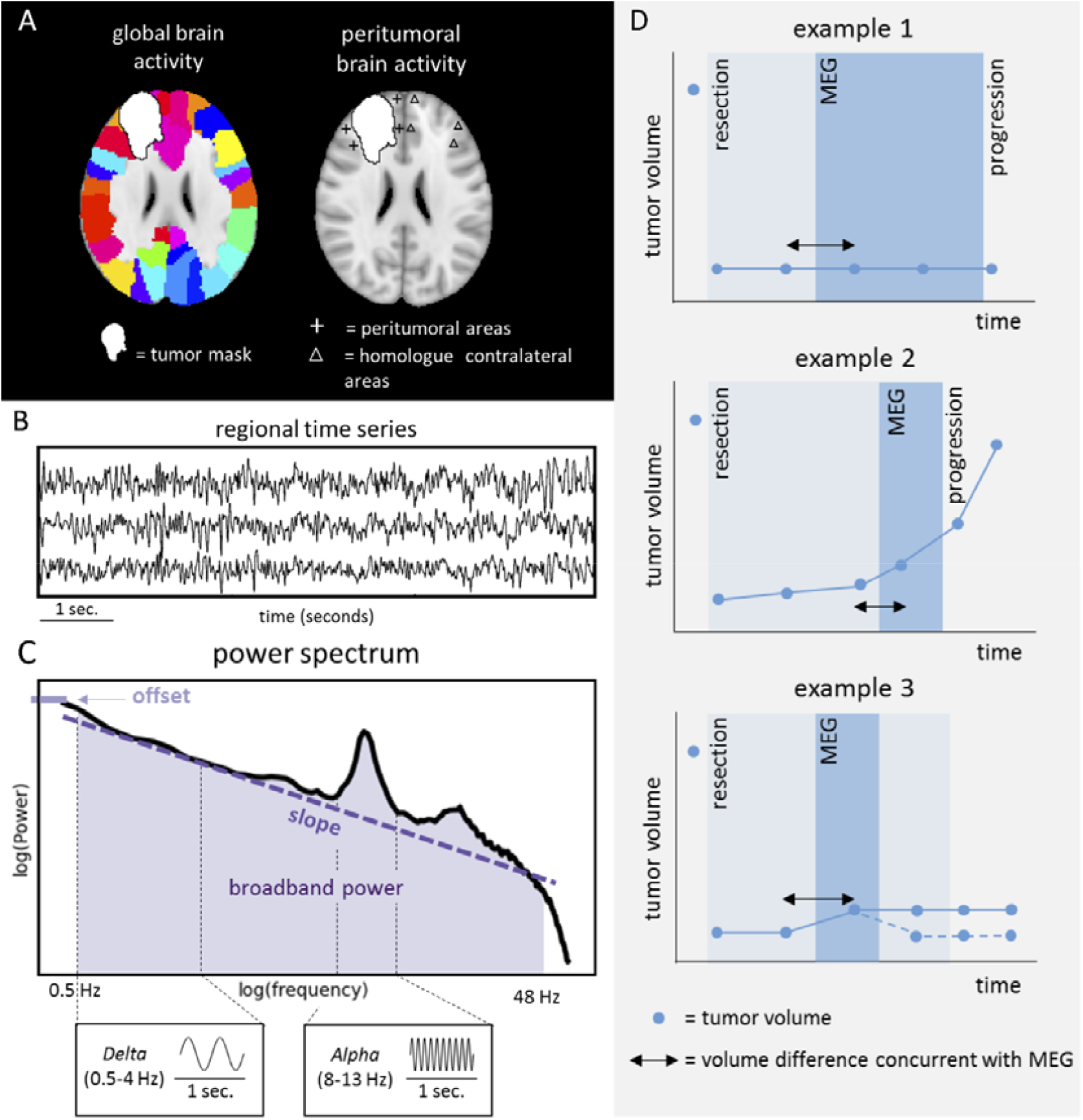
Schematic analysis pipeline of brain activity and tumor volumes. A) Time-series were extracted across regions (left panel) and then averaged to obtain estimates of global brain activity. Peritumoral brain activity was obtained using peritumoral areas (cross hairs) at maximum 3 cm of the tumor or resection cavity and thereafter normalized according to activity of homologue contralateral regions (triangles). B) Three exemplar regional time-series. C) The power spectrum shows which frequencies, and with which strength, are present in the time-series, with slower frequencies on the left (e.g. delta 0.5-4Hz) and faster frequencies on the right (e.g. alpha 8-13Hz). Broadband power was defined as the area under the power spectrum between 0.5-48Hz (shaded area). The offset is the power at the lowest included frequency (0.5Hz). The slope of the power spectrum is indicated by the dashed line. In D, three hypothetical trajectories of patients’ radiological volumes, magnetoencephalography (MEG) measurements and progression as determined by the tumor board are shown. Tumor volume was obtained from all available scans (indicated by blue dots). Example 1 represents a hypothetical patient with stable tumor volume. Example 2 represents a hypothetically radiologically progressive patient (i.e. increasing volume). Example 3 represent a hypothetical patient classified as having radiologically stable tumor volume, because the initial small increase in tumor volume became stable at the next time points (continuous line) or returned to a lower volume indicating potential pseudoprogression (dashed line). The time between MEG and progression as defined by the tumor board is marked with shaded blue and may deviate from radiological tumor growth as operationalized in our study.

Broadband and band-specific power have most often been linked to physiological, cognitive, behavioral, and disease states^14^. However, two new measures of brain activity have recently been introduced^14,15^: the offset of the power spectrum potentially may be more specifically related to neuronal spiking rates^15^, while the slope of the power spectrum may reflect the balance between levels of excitation and inhibition of these neuronal populations^15^. We could thus hypothesize that all three measures of brain activity hold promise as markers of radiological tumor growth.

In a retrospective, cross-sectional setting, we evaluated postoperative global and peritumoral brain activity, operationalized as (1) broadband power, (2) offset and (3) slope of the power spectrum, all three based on source-reconstructed MEG recordings. We related these measures to tumor volume changes measured on routine MRI around the MEG. We hypothesized higher broadband power and offset and lower slope in patients showing increasing tumor volumes, as compared to patients with stable tumor volumes. Finally, we compared our findings for MEG to brain activity recorded with EEG (measured simultaneously with MEG), since MEG is costly and unavailable in most hospitals.

## Results

### Patient characteristics

Our MEG system was replaced in 2010, so we used the subsequent measurements as the main cohort, and the preceding recordings as a validation cohort. The main text will focus on the main cohort, while further information on the validation cohort (n = 21, supplementary table 8) is in the supplementary materials.

In this main cohort, postoperative MEG was available in 50 patients. Four patients were excluded due to unavailable MRI follow-up, one patient was excluded because of low MEG quality. In total, 45 patients were included (table 1). Increasing radiological tumor volumes on the two scans surrounding the MEG (see figure 1 above for a schematic representation of our analysis pipeline) were found in 18 patients (40%), of whom 7 (39%) had IDH-wildtype glioma, 8 (44%) had IDH-mutant 1p/19q non-codeleted glioma, 2 (11%) had IDH-mutant 1p/19q codeleted glioma, and 1 (6%) had an unknown molecular subtype.

**Table 1.**
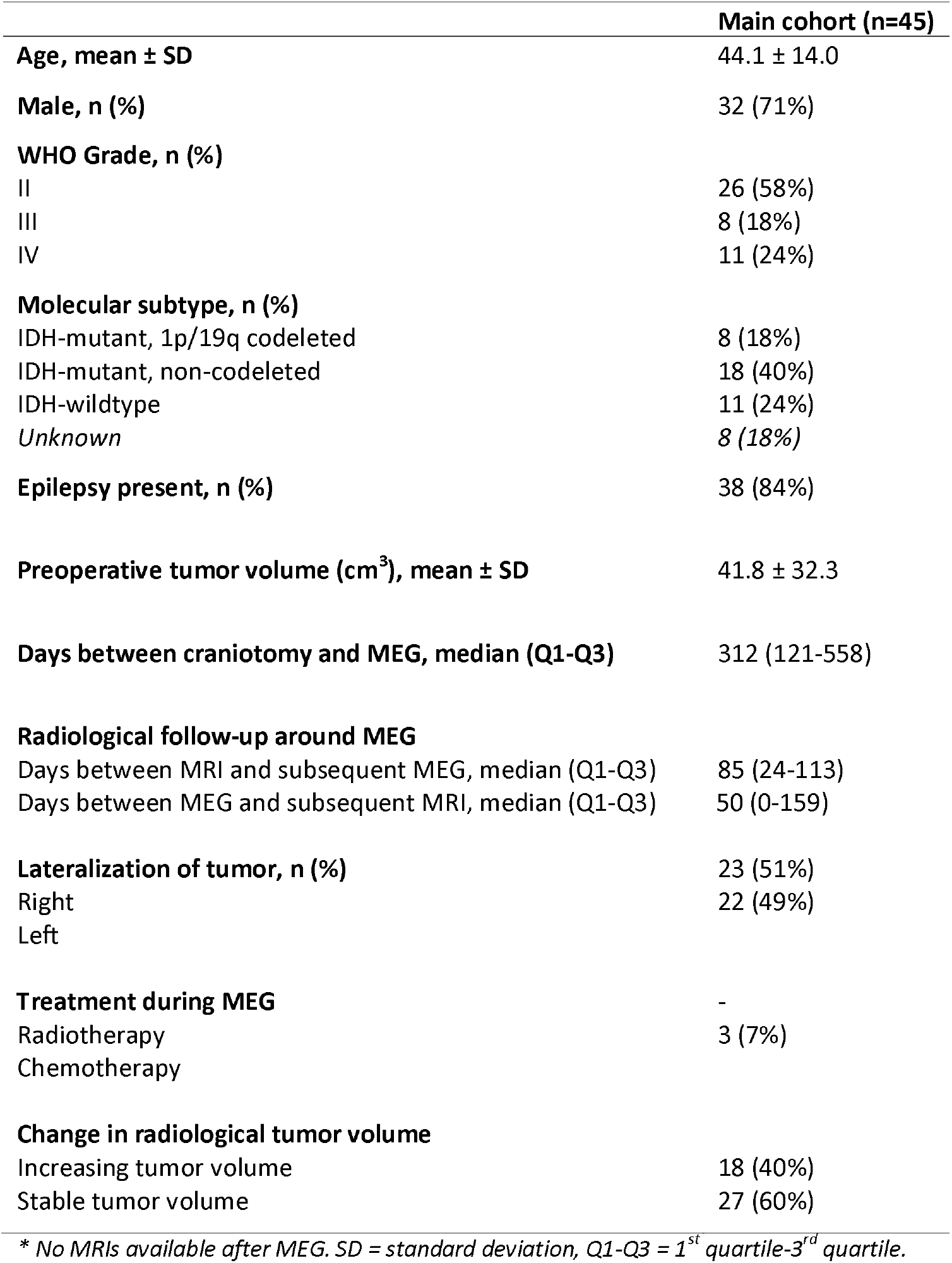
Patient characteristics.

**Table 2:**
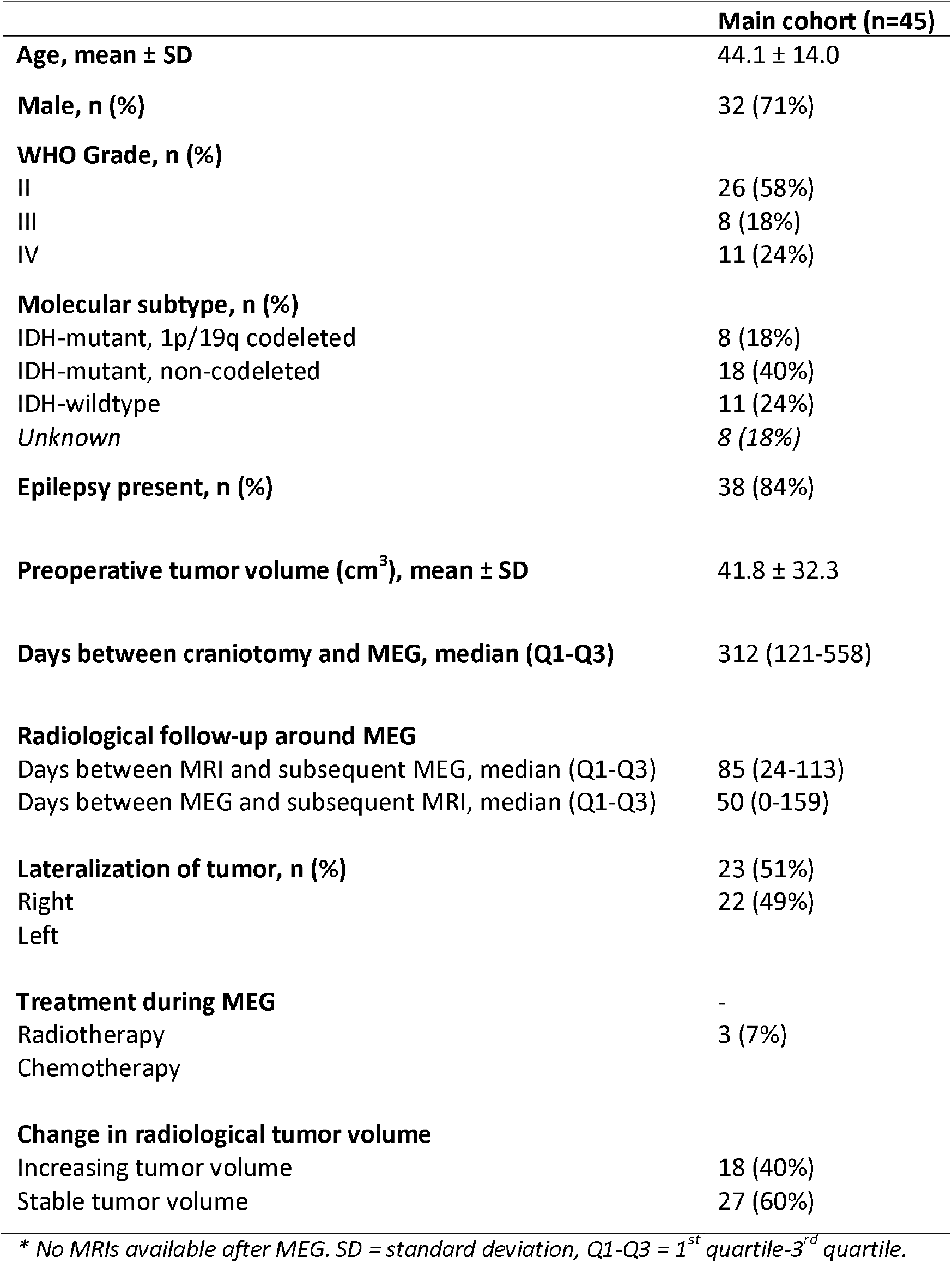
Patient characteristics.

### Global brain activity

For all three measures of source-level brain activity (broadband power, offset and slope of the aperiodic components of the power spectrum), global brain activity did not correlate with tumor volume changes in ml or percentage (figure 2, supplementary table 1, corrected for the 12 tests performed). There were also no differences in brain activity between patients with stable versus increasing tumor volumes (figure 3, supplementary table 2, corrected for the three tests performed). However, global broadband power, offset and slope were significantly higher in both patient groups compared to healthy controls after correction for multiple comparisons (figure 3, supplementary table 2, corrected for the six tests performed).

**Figure 2.**
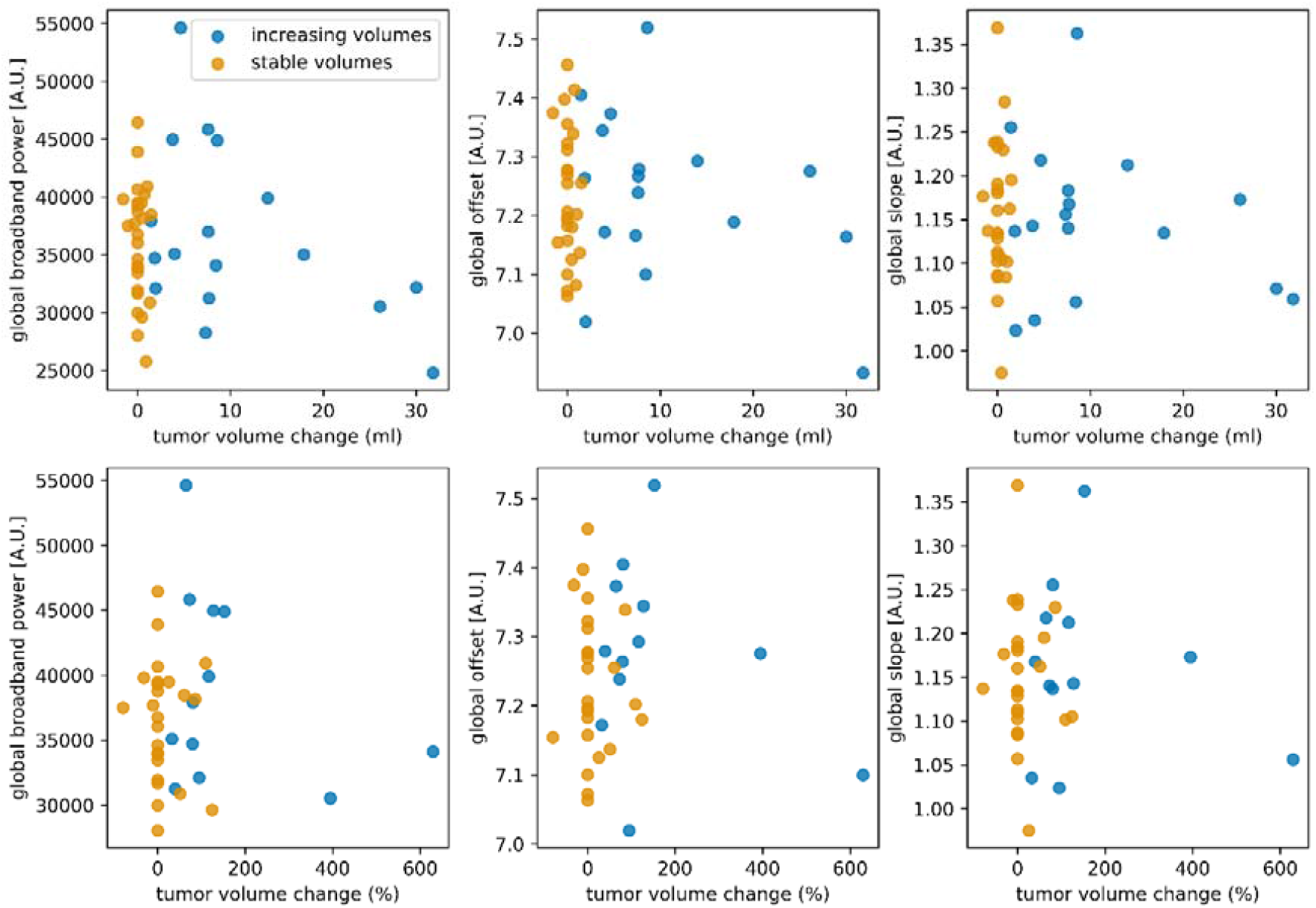
Correlations between tumor volume changes and global brain activity. No significant correlations were found between brain activity and tumor volume change expressed in ml (upper panels) or percent tumor volume change (lower panels) after correction for the three associations between brain activity (broadband power, offset, slope) tested per change score type (ml and percentage). In eight patients, it was not possible to compute percent change, because the initial tumor volume was 0 ml. [A.U.] = arbitrary units.

**Figure 3.**
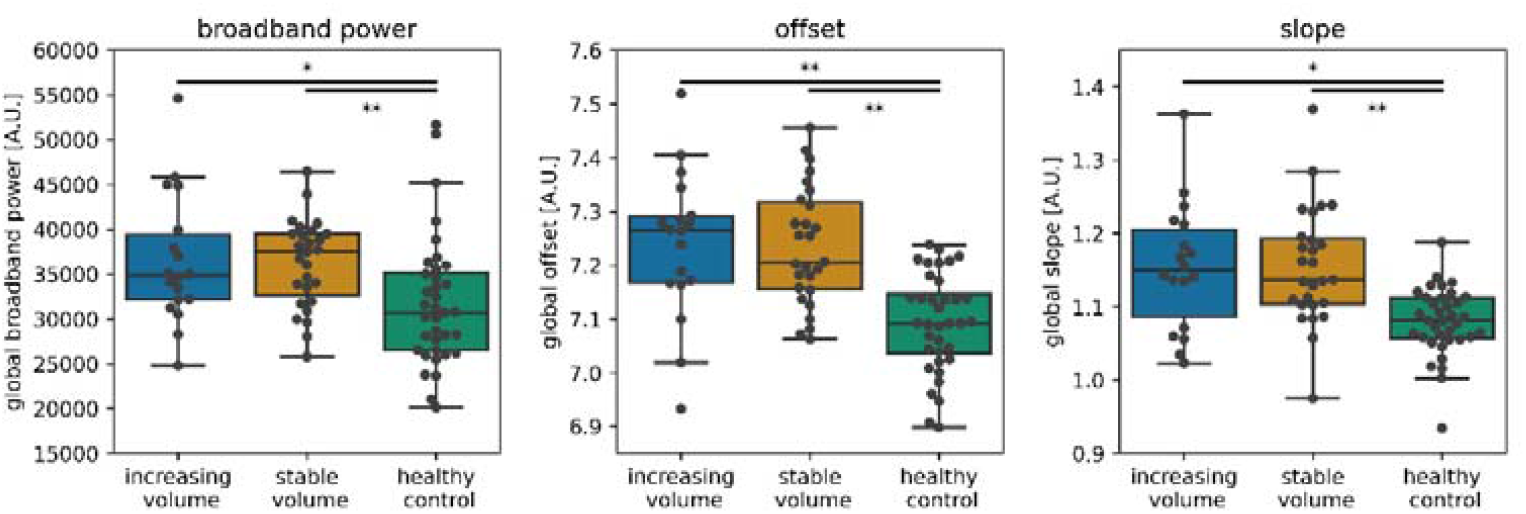
Global brain activity across groups. Global brain activity was not different between patients with stable versus increasing tumor volumes after correcting for the three comparisons drawn. Both glioma groups showed higher global brain activity compared to healthy controls. Individual patients are represented by black dots. * P = 0.001, ** P < 0.001, significant after correction for the six comparisons drawn (three activity values, two group comparisons per activity value). [A.U.] = arbitrary units.

The validation cohort corroborated all main results (supplementary materials, including supplementary figure 7, supplementary table 9, supplementary table 10).

Global broadband power, offset and slope were not related to tumor grade, molecular subtype or presence of epilepsy (supplementary figures 1-3). Preoperative tumor volume was not significantly related to global broadband power (rho(43) = 0.29, *P* = 0.051), however significant correlations with global offset and slope were found (rho(43) = 0.38, *P* = 0.010 and rho(43) = 0.37, *P* = 0.013, respectively, supplementary figure 4).

### Peritumoral brain activity

Peritumoral brain activity (absolute and normalized for homologue contralateral brain activity to account for preexistent regional differences in activity) did not correlate with absolute or percentage tumor growth after correction for the six comparisons drawn per brain activity measure and change score (supplementary table 3), nor did it differ between patients with stable versus increasing radiological tumor volumes after correction for the three tests performed per brain activity type (peritumoral and normalized peritumoral brain activity, supplementary table 4).

However, significantly higher broadband power was found in the peritumoral areas as compared to their contralateral homologues after correction for the three tests performed (W = 108, *P* < 0.001, effect size D = 0.85; figure 4). Similar results were found for slope (W = 45, *P* < 0.001, D = 1.27) and offset (W = 13, *P* < 0.001, D = 1.23). These findings were replicated in the validation cohort (supplementary materials, including supplementary figure 6). Moreover, these differences remained significant in subgroups of patients with either short or long Euclidean distance between their peritumoral and contralateral homologue areas, without correction for multiple comparisons(supplementary table 5).

**Figure 4.**
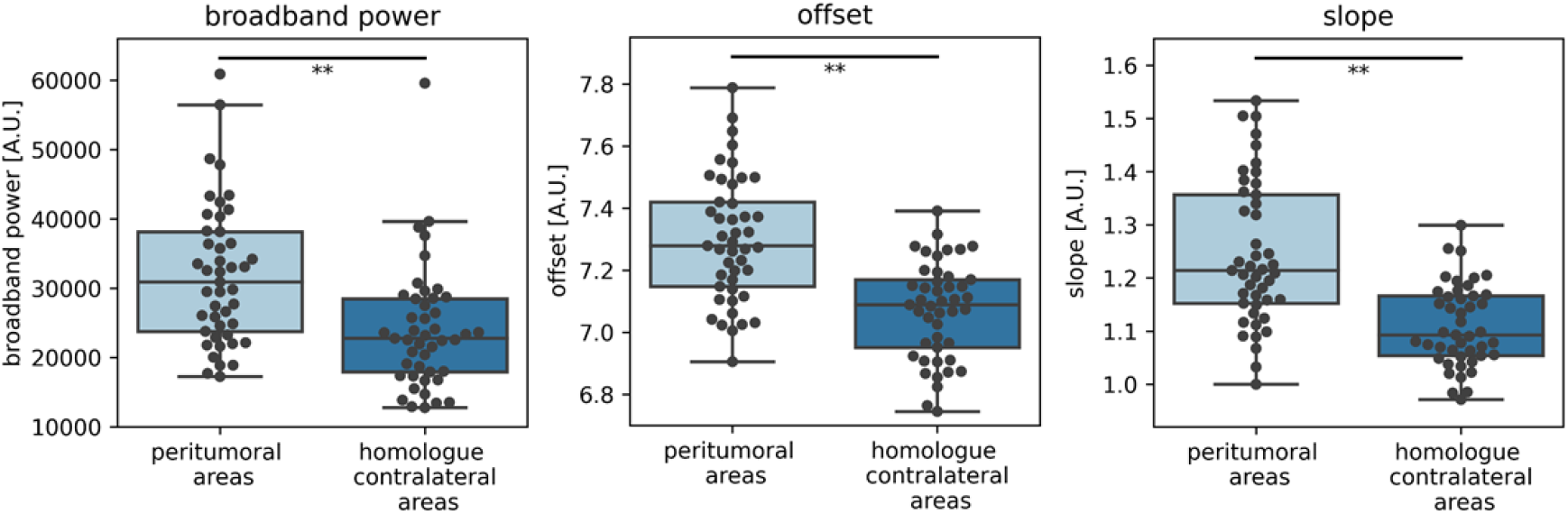
Brain activity of peritumoral and homologue contralateral areas. Peritumoral brain activity was significantly higher compared to the homologue contralateral areas. ** P < 0.001 after correction for the three comparisons (broadband power, offset, slope). [A.U.] = arbitrary units.

### MEG versus EEG

Another cohort of postoperative diffuse glioma patients was prospectively included between 2019-2020 and underwent simultaneous MEG and EEG (MEEG; n = 16; further information on this cohort is in the supplementary materials, including supplementary table 11). In order to explore the potential of sensor-level EEG of recapitulating the significant differences in source-level MEG brain activity between the tumor and contralateral hemispheres, we repeated this analysis for MEG and EEG. Indeed, average brain activity of the tumor hemisphere was generally higher than contralateral brain activity for both modalities (supplementary figure 8).

### Brain activity and time to progression

Progression as assessed by the tumor board was present in 26 patients (58%). Multivariate Cox proportional hazards analyses showed a hazard ratio of 2.48 (95% confidence interval (CI) 1.01-6.07, *P* = 0.048, not corrected for multiple comparisons) for normalized broadband power in relation to time to progression, while slope and offset z-scores were not associated with time to progression (supplementary table 6 and supplementary figure 5). In the validation cohort, there was no relation between brain activity and time to progression (broadband power: HR 1.00 (95% CI 1.00-1.00), *P* = 0.390, supplementary table 7).

## Discussion

Contrary to our hypothesis, global and peritumoral brain activity did not correlate with tumor growth and did not distinguish between patients with stable versus increasing radiological tumor volumes in this heterogeneous cohort of diffuse glioma patients. However, brain activity was significantly higher in patients than in healthy controls. Moreover, patients’ peritumoral brain activity was significantly higher than homologue contralateral areas. Finally, our preliminary findings suggest that EEG is able to recapitulate these increases in tumor-related MEG brain activity.

There are several possible explanations for the lack of association between brain activity and radiological tumor growth. Firstly, our patient cohort was heterogeneous in terms of tumor grade and molecular subtype, which may have obscured more subtle differences in brain activity relating to radiological tumor growth, particularly since histopathological subgroup differences in brain activity have been reported^16,17^. Patients were also in different phases of the disease and their treatment. It is unclear what the potentially confounding effects of chemotherapy and/or radiotherapy are on brain activity measurements with MEG and EEG.

Secondly, most of our patients had epilepsy and used anti-epileptic drugs (AEDs), but the relationship between activity-dependent tumor growth and epilepsy is unknown. In light of the bidirectional relationship between neuronal activity and glioma growth, one could hypothesize that suppressing neuronal activity with AEDs might reduce tumor growth^7,8,12,18^. Indeed, higher seizure frequency is related to worse prognosis^19^. Epidemiological studies have revealed contrasting relationships between AED use and (progression-free) survival: some report longer survival in patients on AEDs^20-24^, others do not^25,26^. In this relatively small and heterogeneous sample, we did not find significant differences in brain activity between patients with or without epilepsy.

Thirdly, determining tumor volumes on MRI and subsequently classifying volume changes as stable or increasing is challenging and might be unreliable depending on imaging techniques used, although MRI currently remains the best marker for progression. In this retrospective study, inclusion of older data meant that additional and potentially informative MRI sequences, e.g. perfusion imaging, were not available in all patients. Therefore, subtle tumor growth might have gone unnoticed, while areas may also have been classified incorrectly as tumor due to pseudoprogression^27,28^, particularly in HGG where tumor volumes were only determined on T1-weighted images after contrast injection. We did mitigate incorrect classifications as much as possible by evaluating all available scans, also those acquired after MEG.

Finally, our indirect measure of neuronal activity may be the culprit for a lack of effect: the animal studies reporting a bidirectional interaction between neuronal activity and glioma growth directly measured neuronal activity^5^. Conversely, broadband power, offset and slope based on MEG are indirect proxies of neuronal activity^15,29^. Little is known on these proxies in terms of spatial sensitivity and lateralization effects, which may have impacted the lack of findings in our cohort. Moreover, we limited our analysis to the broadband frequency range up to 48Hz and excluded the potentially interesting higher gamma frequency range from our analyses, since it reflects only a very small part of the spectrum and shows less consistent correlations with neuronal spiking^29^. Future studies may explore the relevance of specific frequency ranges in relation to tumor growth. At the same time, our findings of higher broadband power and offset in the peritumoral regions do corroborate previous clinical findings using electrocorticography in GBM patients^7^. It could therefore also be that cross-sectionally measured brain activity is not able to differentiate between radiological progression and stable disease. To definitively assess whether brain activity is useful as a marker of tumor progression, future prospective, longitudinal studies in more homogeneous glioma populations and with more extensive standardized MRI and clinical evaluations are necessary.

Such studies may benefit from our findings: broadband power and offset were higher in patients than controls, and were higher in the peritumoral areas and/or ipsilateral hemisphere as compared to the contralateral hemisphere, for both MEG and EEG. In contrast to our expectations, the slope was steeper in patients than in controls and was higher in peritumoral areas, which potentially indicates relatively lower excitation than inhibition^14,15^. More shallow slopes were expected in patients and particularly in the peritumoral areas, since glioma has glutamate-dependent neurogliomal synapses that may raise the excitation-inhibition balance^7^. However, knowledge on the excitation-inhibition balance *in vivo* is particularly scarce. Taken together, these findings suggest that focusing on brain activity of the peritumoral areas (as compared to their contralateral homologue areas) may potentially increase sensitivity to tumor growth when longitudinally following patients. Furthermore, our results may indicate that larger patient cohorts could also be investigated with EEG instead of MEG.Tthe spectral slope does not seem a promising measure of brain activity to assess glioma growth. With respect to progression-free survival, our results corroborate previous findings of shorter PFS in patients with higher broadband power^12,13^, but indicate that the aperiodic components of the power spectrum do not associate with PFS. Future studies may further investigate whether periodic components of the power spectrum are indeed particularly sensitive to PFS.

In conclusion, brain activity is higher in glioma patients as compared to controls, and is higher in the peritumoral areas compared to their contralateral homologues across cohorts and MEG/EEG modalities. However, brain activity does not relate to ongoing radiological glioma growth. Longitudinal studies in more homogeneous and larger cohorts may use these insights to further explore the relevance of non-invasively measured brain activity as a marker of tumor progression.

## Methods

### Subjects

Adult patients were selected from a previously collected cohort, included at the Amsterdam University Medical Centers (Amsterdam UMC, location VUmc) between 2007-2018^12,13,30^. Patients had undergone tumor resection and had histopathologically confirmed diffuse glioma World Health Organization grade II, III or IV^31^. Patients were aged 18 years or older, and had no history of psychiatric or neurologic disease. Isocitrate dehydrogenase (IDH) mutation and 1p/19q codeletion status were obtained after 2016^31^. All patients underwent an MEG measurement after tumor resection. Tumor progression at the MEG timepoint was determined by the multidisciplinary tumor board, based on clinical and radiological characteristics. Time to tumor progression from the MEG was obtained in weeks.

For comparison and normalization, we also included 36 healthy controls (HCs) from a previously described cohort^32,33^, matched to patients at the group level for age, sex and educational level (Verhage system^34^).

### Radiological tumor growth

Routine MRIs up to one year after MEG were used for tumor segmentation. MRI was performed every 3 months for high-grade glioma (HGG), and every 6 months for low-grade glioma (LGG). For LGG, masks were created by segmenting the hyperintense area(s) on T2-weighted and/or FLAIR images. For HGG, contrast enhancing areas on T1-weighted images were masked. Tumor masks were semi-automatically segmented (smart brush tool of iPlan v3.0; BrainLAB AG, Feldkrichen, Germany) under supervision of an experienced neuroradiologist [BM]. Total mask volume was calculated for each MRI.

We first assessed radiological tumor growth as two continuous measures, by subtracting tumor volume on the last MRI before MEG from the first MRI after MEG, and calculating both an absolute volume change and a percentage volume change. Secondly, we classified patients into two groups: stable tumor volume if the difference in volume was <1 ml and increasing tumor volume if the volume increase was >5 ml. Tumor volume increases between 1-5 ml were classified as increasing tumor volume if the following MRI showed further tumor growth, otherwise they were classified as stable tumor volume (figure 1D).

### MEG acquisition and analyses

In the main cohort, MEG data was obtained with a 306-channel whole-head system (Elekta Neuromag Oy, Helsinki, Finland) with a sample frequency of 1250 Hz, and an online anti-aliasing (410 Hz) and high-pass (0.1 Hz) filter. Patients were in supine position inside a magnetically shielded room (VacuumSchmelze GmbH, Hanau, Germany) and were instructed to keep their eyes closed and stay awake during the 5 minute recording.

Data was visually inspected and a maximum of 12 malfunctioning channels were excluded. Temporally extended Signal Space Separation in Maxfilter software (Elekta Neuromag Oy, version 2.2.15)^35,36^ was applied to remove artefacts, followed by application of a single-pass finite impulse response filter between 0.5-48 Hz.

Patients’ head positions were recorded with four or five head position coils. The head-localization coil positions were digitized, as well as the outline of the participants’ scalp and nose (∼500 points), using a 3D digitizer (Fastrak, Polhemus, Colchester, VT). Scalp surfaces of all subjects were co-registered to their structural MRIs using an in-house developed surface-matching procedure, with an estimated resulting accuracy of 4 mm^37^. The Automated Anatomical Labelling (AAL^38^) atlas was used to define 78 cortical regions of interest (ROIs). Broadband (0.5-48Hz) time-series were estimated for the centroid of each of these ROIs by using an atlas-based beamforming approach described previously^39-41^ (and see figure 1A (left panel) and 1B). Specifically, an equivalent current dipole was used as source model, and a single sphere, which was fitted to the outline of the scalp as obtained from the co-registered MRI, was used as a volume conductor model. A scalar beamformer implementation (beamformer, version 2.1.28; Elekta Neuromag Oy) similar to Synthetic Aperture Magnetometry^41^ was used to compute broadband beamformer weights, which were subsequently normalized^42^. Broadband data were used for the computation of the beamformer weights, singular value truncation (with the default setting of 1e-06 times the maximum singular value) was used when inverting the data covariance matrix to deal with the rank deficiency of the data after SSS (∼70 components), and a unity noise covariance matrix was used for the estimation of the optimum source orientation using Singular Value Decomposition^43^.

In order to assess representative peritumoral brain activity, ten locations were manually selected in the grey matter around the tumor and/or resection cavity, in patients’ native MRI space. In doing so, the potential shift of brain tissue due to the tumor and/or resection cavity was taken into account. These locations were all within 3 cm distance of the rim of the tumor mask and were placed in areas without evident edema. After establishing these ten locations, their ten homologue contralateral areas were also selected for analysis (figure 1A right panel). The average Euclidean distance between the peritumoral and contralateral regions was determined for each patient. Scalar beamforming was performed to extract peritumoral and homologue contralateral time-series.

Time-series were split into epochs of 13.1s and visually inspected. The first ten artifact-free epochs were selected for further analysis. To assess the robustness of results with different epoch selection, the main analyses were repeated on the ten epochs with the highest alpha peak (maximum power in the 4-13 Hz band), since higher alpha power has been reported to correlate with less subjective sleepiness^44^. The presence of a clear alpha peak reduces the probability that the subject was drowsy, which may impact brain activity^45^. This alternative epoch selection resulted in identical non-significant results regarding brain activity and tumor volume changes (see supplementary materials), so further analyses were performed on the first ten epochs.

### Global brain activity

Brain activity was assessed using (1) absolute broadband power, and (2) offset and (3) slope of the power spectrum. For each epoch and each brain region, the power spectrum was obtained (figure 1C) based on the signal measured in arbitrary units due to the beamformer implementation, using Welch’s method with a Hamming window, and subsequently averaged across all epochs per patient. Broadband power was then calculated as the area under the curve between 0.5-48 Hz and averaged across all regions to obtain a single value per patient.

The offset and the slope of the power spectra were calculated using the Fitting Oscillations & One Over F (FOOOF) toolbox (https://github.com/fooof-tools/fooof) implemented in Python^14^. The non-oscillatory part of individual power spectra were fitted with an exponential function L: L = b – log(k + F^χ^), where b is the offset, χ is the slope, F represents a vector with the frequencies and k is the knee parameter to set the bending of the aperiodic signal to 0 (i.e. no bending). The offset and slope (figure 1C) were averaged across all regions to obtain a single value per patient.

### Peritumoral brain activity

Broadband power, slope and offset were also calculated with time-series from the selected peritumoral areas and their contralateral homologues. To account for intraindividual variations due to the different tumor locations, normalized peritumoral brain activity was calculated by taking the ratio between brain activity of the peritumoral areas and their homologues in the contralateral hemisphere (figure 1A right panel).

### Statistical analysis

Differences in demographics between patients and HCs were assessed using a Students’ t-test (age) and χ^2^-tests (sex, education).

In order to test our main hypotheses, brain activity was correlated with tumor growth using Spearman’s correlations, and compared between stable versus increasing volume groups with Mann-Whitney U-tests.

Within patients, peritumoral brain activity was compared to homologue contralateral brain activity using paired Wilcoxon signed-rank tests. These analyses were repeated within subgroups of patients with long (>75^th^ percentile) and short (<25^th^ percentile) average distance between the peritumoral and contralateral homologue regions. Furthermore, brain activity was also compared between tumor grades, molecular tumor subtypes and epilepsy status (present or not) using Kruskal-Wallis tests, and correlated with preoperative tumor volumes using Spearman’s correlation.

We also investigated the relationship of global brain activity with time to progression after MEG using multivariate Cox proportional hazards models with age and tumor grade as covariates, to enable comparison with our previous studies^12,13^. To do so, we computed z-scores of global brain activity based on the means and SDs of HCs. Kaplan-Meier curves were created using median splits of brain activity. Patients without progression as determined by the tumor board were censored after last contact with their treating neuro-oncologist.

Statistical analyses were performed using Python 3.6 (Python Software Foundation, www.python.org) and SPSS version 26 (IBM Corp., Armonk, NY, USA). *P*-values < 0.05 were considered significant, after correcting significance levels for the number of tests performed per analysis using the false discovery rate. In case of significant differences between groups, effect sizes were assessed using Cohen’s D.

## Data Availability

The datasets generated during and/or analyzed during the current study are available from the corresponding author on reasonable request. Our final code will be made available through GitHub to allow broader use, although we note that we made use of already publicly available code for all main analyses.

## Author contributions

All authors contributed to the study conception and design. Material preparation, data collection and analysis were performed by T.N., S.D.K., B.M., J.C.R., C.J.S., P.C.d.W.H., J.D., A.M.E.B., M.E.v.L., P.W., M.C.M.K., M.K., T.W., F.B., J.J.G.G., A.H., L.D.. The first draft of the manuscript was written by T.N. and L.D. and all authors commented on previous versions of the manuscript. All authors read and approved the submitted manuscript, and have agreed both to be personally accountable for the author’s own contributions and to ensure that questions related to the accuracy or integrity of any part of the work, even ones in which the author was not personally involved, are appropriately investigated, resolved, and the resolution documented in the literature.

## Additional information

### Ethics approval

All procedures performed in studies involving human participants were in accordance with the ethical standards of the institutional and/or national research committee and with the 1964 Helsinki Declaration and its later amendments or comparable ethical standards. The study was approved by the Bioethics Committee of the VU University Medical Center.

### Consent to participate

Informed consent was obtained from all individual participants included in the study.

## Figure legends

**Figure 2. Schematic analysis pipeline of brain activity and tumor volumes**

A) Time-series were extracted across regions (left panel) and then averaged to obtain estimates of global brain activity. Peritumoral brain activity was obtained using peritumoral areas (cross hairs) at maximum 3 cm of the tumor or resection cavity and thereafter normalized according to activity of homologue contralateral regions (triangles). B) Three exemplar regional time-series. C) The power spectrum shows which frequencies, and with which strength, are present in the time-series, with slower frequencies on the left (e.g. delta 0.5-4Hz) and faster frequencies on the right (e.g. alpha 8-13Hz). Broadband power was defined as the area under the power spectrum between 0.5-48Hz (shaded area). The offset is the power at the lowest included frequency (0.5Hz). The slope of the power spectrum is indicated by the dashed line. In D, three hypothetical trajectories of patients’ radiological volumes, magnetoencephalography (MEG) measurements and progression as determined by the tumor board are shown. Tumor volume was obtained from all available scans (indicated by blue dots). Example 1 represents a hypothetical patient with stable tumor volume. Example 2 represents a hypothetically radiologically progressive patient (i.e. increasing volume). Example 3 represent a hypothetical patient classified as having radiologically stable tumor volume, because the initial small increase in tumor volume became stable at the next time points (continuous line) or returned to a lower volume indicating potential pseudoprogression (dashed line). The time between MEG and progression as defined by the tumor board is marked with shaded blue and may deviate from radiological tumor growth as operationalized in our study.

**Figure 2. Correlations between tumor volume changes and global brain activity**

No correlations were found between brain activity and tumor volume change expressed in ml (upper panels) or percent tumor volume change (lower panels). In eight patients, it was not possible to compute percent change, because the initial tumor volume was 0 ml. [A.U.] = arbitrary units.

**Figure 3. Global brain activity across groups**

Global brain activity was not different between patients with stable versus increasing tumor volumes. Both glioma groups showed higher global brain activity compared to healthy controls. Individual patients are represented by black dots. * *P* < 0.05, ** *P* < 0.01 after correction for multiple comparisons. [A.U.] = arbitrary units.

**Figure 4. Brain activity of peritumoral and homologue contralateral areas**

Peritumoral brain activity was significantly higher compared to the homologue contralateral areas. ** *P* < 0.01 after correction for multiple comparisons. [A.U.] = arbitrary units.

## References

1 Thust, S. C. et al. Glioma imaging in Europe: A survey of 220 centres and recommendations for best clinical practice. Eur Radiol 28, 3306–3317, doi:10.1007/s00330-018-5314-5 (2018).

2 Villanueva-Meyer, J. E., Mabray, M. C. & Cha, S. Current Clinical Brain Tumor Imaging. Neurosurgery 81, 397–415, doi:10.1093/neuros/nyx103 (2017).

3 Brown, P. D. et al. Detrimental effects of tumor progression on cognitive function of patients with high-grade glioma. J Clin Oncol 24, 5427–5433, doi:10.1200/JCO.2006.08.5605 (2006).

4 Meyers, C. A. & Hess, K. R. Multifaceted end points in brain tumor clinical trials: cognitive deterioration precedes MRI progression. Neuro Oncol 5, 89–95, doi:10.1093/neuonc/5.2.89 (2003).

5 Venkatesh, H. S. et al. Neuronal Activity Promotes Glioma Growth through Neuroligin-3 Secretion. Cell 161, 803–816, doi:10.1016/j.cell.2015.04.012 (2015).

6 Venkatesh, H. S. et al. Targeting neuronal activity-regulated neuroligin-3 dependency in high-grade glioma. Nature 549, 533–537, doi:10.1038/nature24014 (2017).

7 Venkataramani, V. et al. Glutamatergic synaptic input to glioma cells drives brain tumour progression. Nature 573, 532–538, doi:10.1038/s41586-019-1564-x (2019).

8 Venkatesh, H. S. et al. Electrical and synaptic integration of glioma into neural circuits. Nature 573, 539–545, doi:10.1038/s41586-019-1563-y (2019).

9 Murakami, S. & Okada, Y. Contributions of principal neocortical neurons to magnetoencephalography and electroencephalography signals. J Physiol 575, 925–936, doi:10.1113/jphysiol.2006.105379 (2006).

10 Barnes, G. R., Hillebrand, A., Fawcett, I. P. & Singh, K. D. Realistic spatial sampling for MEG beamformer images. Hum Brain Mapp 23, 120–127, doi:10.1002/hbm.20047 (2004).

11 Troebinger, L., Lopez, J. D., Lutti, A., Bestmann, S. & Barnes, G. Discrimination of cortical laminae using MEG. Neuroimage 102 Pt 2, 885–893, doi:10.1016/j.neuroimage.2014.07.015 (2014).

12 Belgers, V. et al. Postoperative oscillatory brain activity as an add-on prognostic marker in diffuse glioma. J Neurooncol 147, 49–58, doi:10.1007/s11060-019-03386-7 (2020).

13 Derks, J. et al. Oscillatory brain activity associates with neuroligin-3 expression and predicts progression free survival in patients with diffuse glioma. J Neurooncol, doi:10.1007/s11060-018-2967-5 (2018).

14 Donoghue, T. et al. Parameterizing neural power spectra into periodic and aperiodic components. Nature Neuroscience 23, 1655–1665 (2020).

15 Gao, R., Peterson, E. J. & Voytek, B. Inferring synaptic excitation/inhibition balance from field potentials. Neuroimage 158, 70–78, doi:10.1016/j.neuroimage.2017.06.078 (2017).

16 de Jongh, A. et al. The influence of brain tumor treatment on pathological delta activity in MEG. Neuroimage 20, 2291–2301, doi:10.1016/j.neuroimage.2003.07.030 (2003).

17 Wilson, T. W., Heinrichs-Graham, E. & Aizenberg, M. R. Potential role for magnetoencephalography in distinguishing low- and high-grade gliomas: a preliminary study with histopathological confirmation. Neuro Oncol 14, 624–630, doi:10.1093/neuonc/nos064 (2012).

18 Venkataramani, V., Tanev, D. I., Kuner, T., Wick, W. & Winkler, F. Synaptic Input to Brain Tumors: Clinical Implications. Neuro Oncol, doi:10.1093/neuonc/noaa158 (2020).

19 Vecht, C. J., Kerkhof, M. & Duran-Pena, A. Seizure prognosis in brain tumors: new insights and evidence-based management. Oncologist 19, 751–759, doi:10.1634/theoncologist.2014-0060 (2014).

20 Lu, V. M., Jue, T. R., Phan, K. & McDonald, K. L. Quantifying the prognostic significance in glioblastoma of seizure history at initial presentation: A systematic review and meta-analysis. Clin Neurol Neurosurg 164, 75–80, doi:10.1016/j.clineuro.2017.11.015 (2018).

21 Toledo, M. et al. Epileptic features and survival in glioblastomas presenting with seizures. Epilepsy Res 130, 1–6, doi:10.1016/j.eplepsyres.2016.12.013 (2017).

22 Pallud, J. & McKhann, G. M. Diffuse Low-Grade Glioma-Related Epilepsy. Neurosurg Clin N Am 30, 43–54, doi:10.1016/j.nec.2018.09.001 (2019).

23 Roh, T. H. et al. Association between survival and levetiracetam use in glioblastoma patients treated with temozolomide chemoradiotherapy. Sci Rep 10, 10783, doi:10.1038/s41598-020-67697-w (2020).

24 Brodie, S. A. & Brandes, J. C. Could valproic acid be an effective anticancer agent? The evidence so far. Expert Rev Anticancer Ther 14, 1097–1100, doi:10.1586/14737140.2014.940329 (2014).

25 van Breemen, M. S. et al. Efficacy of anti-epileptic drugs in patients with gliomas and seizures. J Neurol 256, 1519–1526, doi:10.1007/s00415-009-5156-9 (2009).

26 Berendsen, S. et al. Prognostic relevance of epilepsy at presentation in glioblastoma patients. Neuro Oncol 18, 700–706, doi:10.1093/neuonc/nov238 (2016).

27 Thust, S. C., van den Bent, M. J. & Smits, M. Pseudoprogression of brain tumors. J Magn Reson Imaging 48, 571–589, doi:10.1002/jmri.26171 (2018).

28 van West, S. E. et al. Incidence of pseudoprogression in low-grade gliomas treated with radiotherapy. Neuro Oncol 19, 719–725, doi:10.1093/neuonc/now194 (2017).

29 Manning, J. R., Jacobs, J., Fried, I. & Kahana, M. J. Broadband shifts in local field potential power spectra are correlated with single-neuron spiking in humans. J Neurosci 29, 13613–13620, doi:10.1523/JNEUROSCI.2041-09.2009 (2009).

30 Derks, J. et al. Understanding cognitive functioning in glioma patients: The relevance of IDH-mutation status and functional connectivity. Brain Behav 9, e01204, doi:10.1002/brb3.1204 (2019).

31 Louis, D. N. et al. The 2016 World Health Organization Classification of Tumors of the Central Nervous System: a summary. Acta Neuropathol 131, 803–820, doi:10.1007/s00401-016-1545-1 (2016).

32 Tewarie, P. et al. Structural degree predicts functional network connectivity: a multimodal resting-state fMRI and MEG study. Neuroimage 97, 296–307, doi:10.1016/j.neuroimage.2014.04.038 (2014).

33 Tewarie, P. et al. Functional brain networks: linking thalamic atrophy to clinical disability in multiple sclerosis, a multimodal fMRI and MEG study. Hum Brain Mapp 36, 603–618, doi:10.1002/hbm.22650 (2015).

34 Verhage, F. Intelligentie en leeftijd: Onderzoek bij Nederlanders van twaalf tot zevenenzeventig jaar., (1964).

35 Taulu, S. & Hari, R. Removal of magnetoencephalographic artifacts with temporal signal-space separation: demonstration with single-trial auditory-evoked responses. Hum Brain Mapp 30, 1524–1534, doi:10.1002/hbm.20627 (2009).

36 Taulu, S. & Simola, J. Spatiotemporal signal space separation method for rejecting nearby interference in MEG measurements. Phys Med Biol 51, 1759–1768, doi:10.1088/0031-9155/51/7/008 (2006).

37 Whalen, C., Maclin, E. L., Fabiani, M. & Gratton, G. Validation of a method for coregistering scalp recording locations with 3D structural MR images. Hum Brain Mapp 29, 1288–1301, doi:10.1002/hbm.20465 (2008).

38 Tzourio-Mazoyer, N. et al. Automated anatomical labeling of activations in SPM using a macroscopic anatomical parcellation of the MNI MRI single-subject brain. Neuroimage 15, 273–289, doi:10.1006/nimg.2001.0978 (2002).

39 Hillebrand, A., Barnes, G. R., Bosboom, J. L., Berendse, H. W. & Stam, C. J. Frequency-dependent functional connectivity within resting-state networks: an atlas-based MEG beamformer solution. Neuroimage 59, 3909–3921, doi:10.1016/j.neuroimage.2011.11.005 (2012).

40 Hillebrand, A. et al. Direction of information flow in large-scale resting-state networks is frequency-dependent. Proceedings of the National Academy of Science USA 113, 3867–3872 (2016).

41 Robinson, S. E. Functional neuroimaging by Synthetic Aperture Magnetometry (SAM). Recent Adv Biomagnet, 302–305 (1999).

42 Cheyne, D., Bakhtazad, L. & Gaetz, W. Spatiotemporal mapping of cortical activity accompanying voluntary movements using an event-related beamforming approach. Hum Brain Mapp 27, 213–229, doi:10.1002/hbm.20178 (2006).

43 Sekihara, K., Nagarajan, S. S., Poeppel, D. & Marantz, A. Asymptotic SNR of scalar and vector minimum-variance beamformers for neuromagnetic source reconstruction. IEEE Trans Biomed Eng 51, 1726–1734, doi:10.1109/TBME.2004.827926 (2004).

44 Kaida, K. et al. Validation of the Karolinska sleepiness scale against performance and EEG variables. Clin Neurophysiol 117, 1574–1581, doi:10.1016/j.clinph.2006.03.011 (2006).

45 van Diessen, E. et al. Opportunities and methodological challenges in EEG and MEG resting state functional brain network research. Clin Neurophysiol 126, 1468–1481, doi:10.1016/j.clinph.2014.11.018 (2015).

