## Supplementary materials for "Non-invasively measured brain activity and radiological progression in diffuse glioma"

### Supplementary tables and figures

#### Supplementary table 1: Correlations between tumor volume changes and global brain activity in the main cohort

| | *Correlations with change in tumor volume (ml)* | |
|---|---|---|
| | **All (n=44)** | **Increasing volume (n=17)** |
| **Global broadband power** | Rho=-0.10, P=0.514 | Rho=-0.35, P=0.168 |
| **Global offset** | Rho=-0.09, P=0.567 | Rho=-0.25, P=0.333 |
| **Global slope** | Rho=-0.09, P=0.552 | Rho=-0.04, P=0.881 |
| | *Correlations with change in tumor volume (%)* | |
| | **All (n=37)** | **Increasing volume (n=12)** |
| **Global broadband power** | Rho=0.06, P=0.737 | Rho=-0.18, P=0.572 |
| **Global offset** | Rho=0.03, P=0.876 | Rho=0.06, P=0.846 |
| **Global slope** | Rho=-0.03, P=0.856 | Rho=0.15, P=0.633 |

Note. Significance was determined after correction for the six tests performed per change score (ml or percentage).

#### Supplementary table 2: Global brain activity across groups in the main cohort

| | Increasing volume (n=18) | Stable volume (n=27) | HC (n=36) | Increasing vs. stable volume | Increasing volume vs. HC | Stable volume vs. HC |
|---|---|---|---|---|---|---|
| **Global broadband power** | 34873 (32137-39406) | 37517 (32705-39501) | 30765 (26522-35183) | U=230, P=0.386 | U=195, P=0.009*, D=0.64 | U=260, P=0.001*, D=0.71 |
| **Global offset** | 7.26 (7.17-7.29) | 7.21 (7.16-7.32) | 7.09 (7.04-7.15) | U=232, P=0.404 | U=112, P<0.001*, D=1.24 | U=169, P<0.001*, D=1.41 |
| **Global slope** | 1.15 (1.09-1.21) | 1.14 (1.10-1.19) | 1.08 (1.06-1.11) | U=240, P=0.477 | U=146, P=0.001*, D=1.07 | U=181, P<0.001*, D=1.17 |

Note. Median values (1st quartile-3rd quartile) are reported. HC = healthy control. * = P significant after correction for multiple comparisons, which entailed correction for three tests performed comparing the increasing versus stable volume groups, and six comparisons drawn between increasing volume and stable volume patients versus healthy controls.

**Supplementary table 3: Correlations between tumor volume changes and peritumoral brain activity**

| | Correlations with change in tumor volume (ml) | |
|---|---|---|
| | **All (n=44)** | **Increasing volume (n=17)** |
| ***Peritumoral brain activity*** | | |
| **Broadband power** | rho=-0.08, P=0.641 | rho=0.21, P=0.513 |
| **Offset** | rho=0.05, P=0.776 | rho=0.39, P=0.208 |
| **Slope** | rho=0.18, P=0.273 | rho=0.59, P=0.045 |
| ***Normalized peritumoral brain activity*** | | |
| **Broadband power** | rho=-0.32, P=0.053 | rho=0.41, P=0.183 |
| **Offset** | rho=-0.07, P=0.698 | rho=0.57, P=0.051 |
| **Slope** | rho=0.29, P=0.077 | rho=0.59, P=0.042 |
| | ***Correlations with change in tumor volume (%)*** | |
| | **All (n=37)** | **Increasing volume (n=12)** |
| ***Peritumoral brain activity*** | | |
| **Broadband power** | rho=-0.08, P=0.621 | rho=0.03, P=0.914 |
| **Offset** | rho=0.01, P=0.962 | rho=0.04, P=0.897 |
| **Slope** | rho=0.10, P=0.575 | rho=0.11, P=0.729 |
| ***Normalized peritumoral brain activity*** | | |
| **Broadband power** | rho=-0.39, P=0.018 | rho=-0.06, P=0.863 |
| **Offset** | rho=-0.19, P=0.259 | rho=-0.06, P=0.863 |
| **Slope** | rho=0.16, P=0.332 | rho=-0.06, P=0.846 |

Note. Significance was determined after correction for the six tests performed per change score ml or percentage) and brain activity type (peritumoral or normalized peritumoral).**Supplementary table 4: *Peritumoral* brain activity *across groups***

| | Peritumoral brain activity | | | Normalized peritumoral brain activity | | |
|---|---|---|---|---|---|---|
| | **Increasing volume (n=18)** | **Stable volume (n=27)** | **U-statistic, p-value** | **Increasing volume (n=18)** | **Stable volume (n=27)** | **U-statistic, p-value** |
| **Broadband power** | 26794 (22077-38232) | 32568 (26351-36437) | U=193, P=0.126 | 1.10 (1.04-1.55) | 1.40 (1.06-1.78) | U=191, P=0.116 |
| **Offset** | 7.27 (7.15-7.42) | 7.29 (7.16-7.43) | U=242, P=0.495 | 1.03 (1.01-1.05) | 1.03 (1.01-1.04) | U=227, P=0.360 |
| **Slope** | 1.24 (1.18-1.38) | 1.20 (1.14-1.32) | U=185, P=0.091 | 1.15 (1.06-1.19) | 1.08 (1.03-1.15) | U=189, P=0.108 |

Note. Significance was determined after correction for the three tests performed per brain activity type (peritumoral or normalized peritumoral).

**Supplementary table 5: Distance-independence of differences between peritumoral and contralateral homologue areas**

| | Short distance (n=11) | | | Long distance (n=11) | | |
|---|---|---|---|---|---|---|
| | **Peritumoral** | **Contralateral** | **W-statistic, p-value** | **Peritumoral** | **Contralateral** | **W-statistic, p-value** |
| **Broadband power** | 32569 (23777-35738) | 23202 (22449-29891) | W=8, P=0.026* | 29513 (13437-23357) | 19105 (13437-23357) | W=5, P=0.013* |

| | | | | | | |
|---|---|---|---|---|---|---|
| **Offset** | 7.26 (7.14-7.36) | 7.10 (6.97-7.15) | W=3, P=0.008* | 7.28 (7.05-7.50) | 6.92 (6.87-7.07) | W=1, P=0.004* |
| **Slope** | 1.16 (1.12-1.23) | 1.07 (1.04-1.10) | W<0.001, P=0.003* | 1.22 (1.16-1.40) | 1.05 (1.02-1.17) | W<0.001,P=0.003* |

Note. Median values (1st quartile-3rd quartile) are reported. * = P significant without correction for multiple comparisons.

**Supplementary figure 1: Global brain activity across tumor grades**

*No differences were found between tumor grades with respect to global broadband power ($\chi^2(2)=1.32$, $P=0.518$), offset ($\chi^2(2)=0.66$ $P=0.719$) or slope ($\chi^2(2)=0.01$, $P=0.9930$), without correction for multiple comparisons. [A.U.] = arbitrary units.*

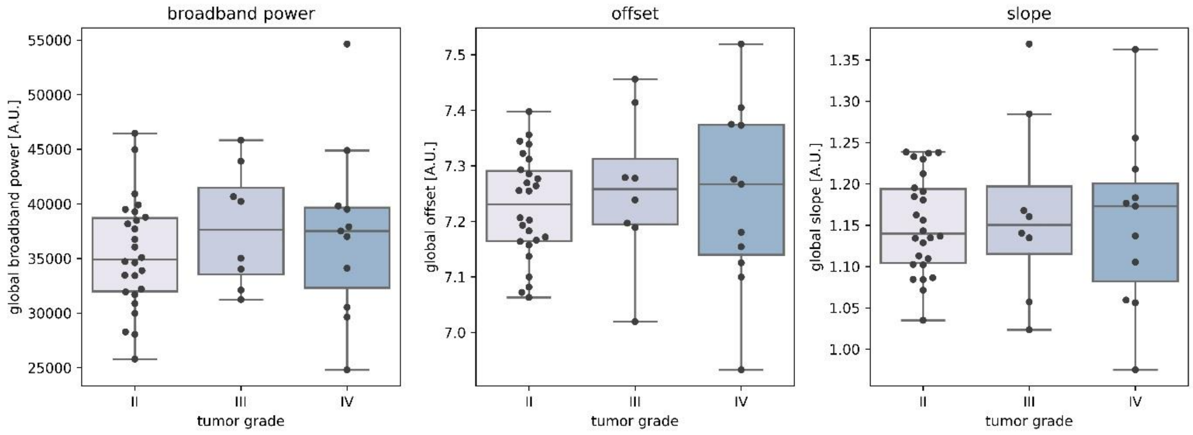


**Supplementary figure 2: Global brain activity across molecular subtypes**

*No differences were found between molecular subtypes with respect to global broadband power ($\chi^2(2)=1.09$, $P=0.579$), offset ($\chi^2(2)=0.01$, $P=0.995$) or slope ($\chi^2(2)=0.01$, $P=0.994$), without correction for multiple comparisons. IDH-mut=Isocitrate dehydrogenase mutant, IDH-wt=Isocitrate dehydrogenase wildtype , codel=codeleted, non-codel=non-codeleted. [A.U.] = arbitrary units.*

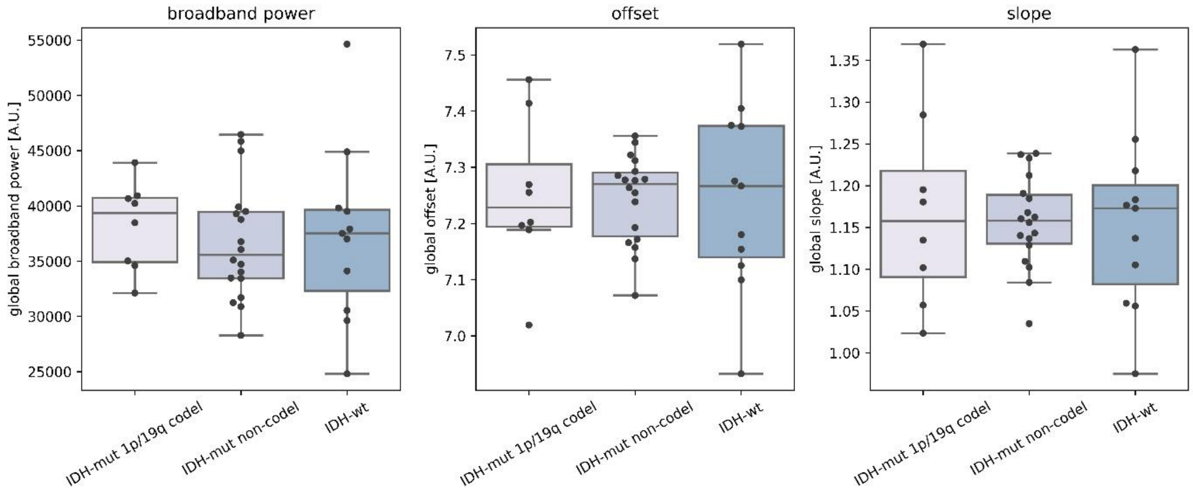

**Supplementary figure 3: Global brain activity according to presence of epilepsy**

*No significant differences between patients with (n=38) and without epilepsy (n=7) were found with respect to global broadband power (U=130, P=0.469), offset (U=91, P=0.097) or slope (U=82, P=0.057), without correction for multiple comparisons. [A.U.] = arbitrary units.*

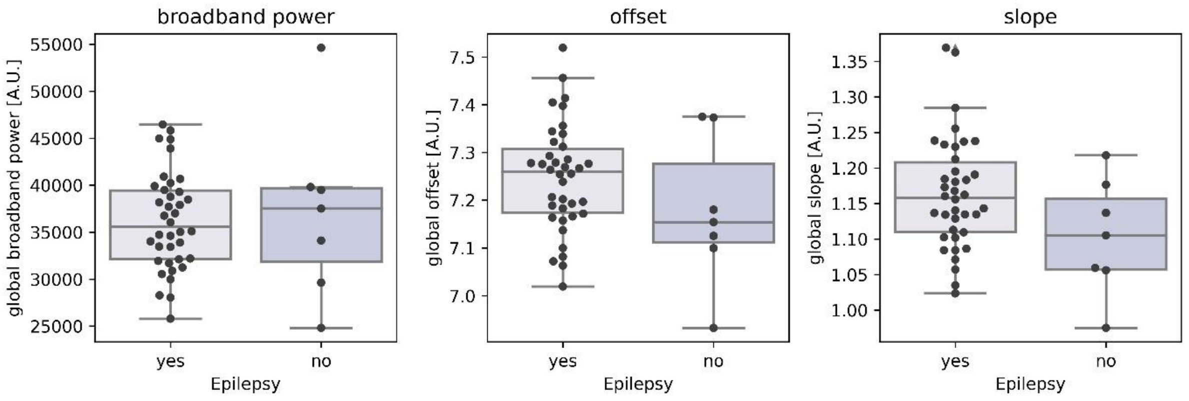


**Supplementary figure 4: Correlations between global brain activity and preoperative tumor volume**

Left *Spearman's rho correlations were not significant between preoperative tumor volume and global broadband power (rho(43)=0.29, P=0.051).* Middle *A significant correlation was found between preoperative tumor volume and the global offset (rho(43)=0.38, P=0.010) and slope (rho(43)=0.37, P=0.013,* Right*). These correlations were not corrected for multiple comparisons. [A.U.] = arbitrary units.*

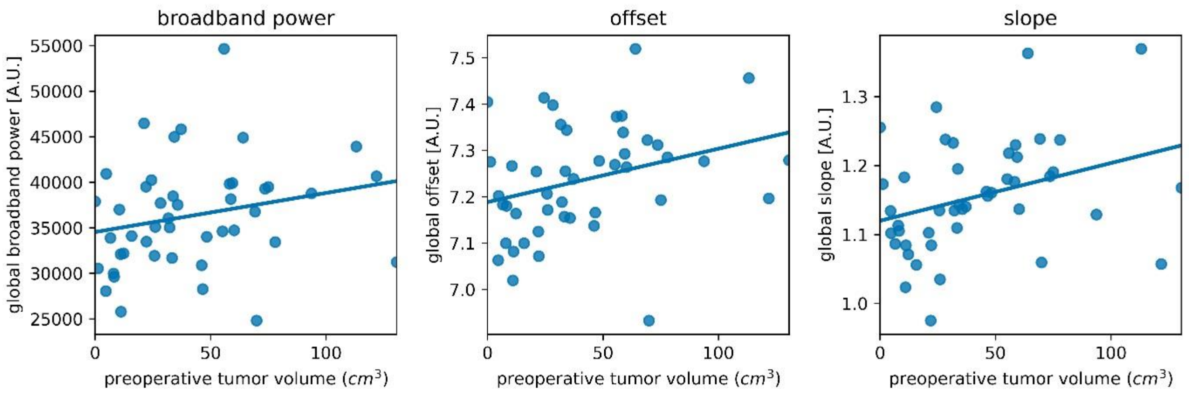


**Supplementary table 6: Results of multivariate Cox regression analysis in the main cohort**

| Covariates | Global broadband power | Global offset | Global slope |
|---|---|---|---|
| **Global brain activity** | 2.48 (1.01-6.07), P=0.048* | 1.43 (0.83-2.47), P=0.192 | 1.17 (0.73-1.88), P=0.516 |
| **Age** | 0.98 (0.95-1.02), P=0.290 | 0.99 (0.96-1.03), P=0.755 | 0.99 (0.96-1.03), P=0.689 |
| **Grade II** | *Reference* | *Reference* | *Reference* |
| **Grade III** | 0.41 (0.09-1.89), P=0.251 | 0.52 (0.12-2.35), P=0.398 | 0.56 (0.13-2.50), P=0.447 |
| **Grade IV** | 5.52 (1.90-16.1), P=0.002* | 5.94 (2.10-16.79), P=0.001* | 6.01 (2.15-16.8), =<0.001* |

Note. Hazard ratios (95% confidence interval) are reported. * = P<0.05 without correction for multiple comparisons.

**Supplementary table 7: Results of multivariate Cox regression analysis in the validation cohort**

| Covariates | Global broadband power | Global offset | Global Slope |
|---|---|---|---|
| **Global brain activity** | 1.00 (1.00-1.00), P=0.390 | 1.73 (0.04-73.69), P=0.774 | 0.80 (0.00-299.61), P=0.942 |
| **Age** | 1.00 (0.93-1.08), P=0.978 | 0.99 (0.92-1.06), P=0.716 | 0.99 (0.20-1.06), P=0.683 |
| **Grade II*** | *Reference* | *Reference* | *Reference* |
| **Grade IV** | 3.98 (1.17-13.57), P=0.027* | 3.77 (1.03-13.87), P=0.046* | 3.44 (1.02-11.63), P=0.047* |

Note. Hazard ratios (95% confidence interval) are reported. Only 3 patients had grade III glioma, which were therefore excluded from this analysis. * = P<0.05 without correction for multiple comparisons.

**Supplementary figure 5: Kaplan-Meier curves**
*These curves visualize the difference between high and low normalized brain activity and their relation with time to progression (in weeks) as established by the tumor board, counting from the day of MEG. Three patients already had progression before MEG and were excluded from this analysis. Groups were dichotomized based on a median split of the brain activity measure (low activity in blue versus high activity in red).*

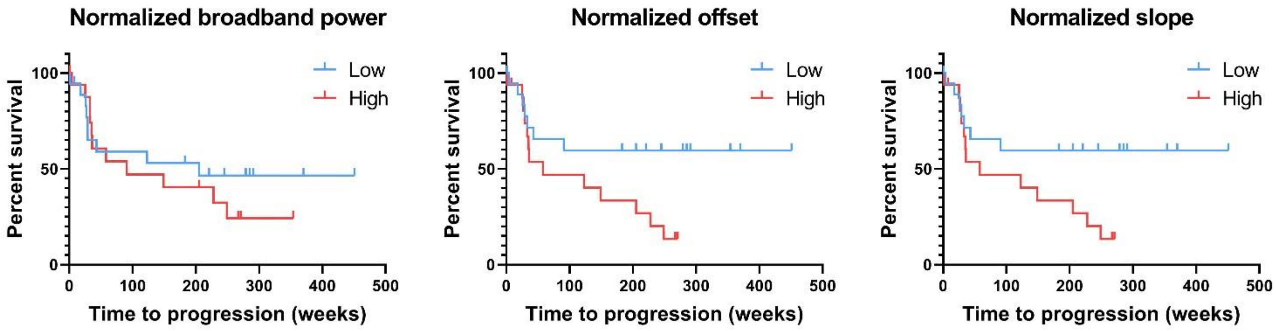


### Supplementary information validation cohort

#### *Data acquisition*

In the validation cohort, MEG recordings were performed in eyes-closed supine position with a 151-channel whole head system (CTF Systems, Inc., Port Coquitlam, BC, Canada) in the same magnetically shielded room as the Elekta system (VacuumSchmelze GmbH, Hanua, Germany). The sample frequency was 625Hz and the recording was bandpass filtered between 0-150Hz. Head position was determined at the beginning and ending of each recording using head position coils placed at the nasion, and left and right pre-auricular points. Data were filtered between 0.5-48Hz and split into epochs of 4096 samples. Twenty artifact-free epochs were selected after visual inspection.

#### *Global and peritumoral brain activity*

Global, peritumoral and contralateral homologue broadband power, offset and slope were calculated identically to the main cohort, and group differences and associations were tested in the same way as well.

***Results***

In total, 21 patients were included, after excluding patients due to direct re-resection after MEG (n=2), and low MEG quality (n=1). The majority of this cohort had an IDH-wildtype diffuse glioma, see supplementary table 7 for patient characteristics. Only 5 patients were classified as having increasing tumor volume, while 16 patients had stable tumor volumes.

Corroborating the findings in the main cohort, there were no associations between brain activity and tumor volume changes (supplementary table 8), and there were no differences in brain activity between groups (supplementary table 9, supplementary figure 7).

Also in line with the main cohort, peritumoral brain activity was significantly higher than contralateral homologue brain activity (supplementary figure 6).

**Supplementary table 8: Patient characteristics of validation cohort**

| | **Validation cohort (n=21)** |
|---|---|
| **Age, mean ± SD** | 48.6 ± 11.3 |
| **Male, n (%)** | 16 (76%) |
| **WHO Grade, n (%)** | |
| II | 8 (38%) |
| III | 3 (14%) |
| IV | 10 (48%) |
| **Molecular subtype, n (%)** | |
| IDH-mutant, 1p/19q codeleted | 3 (14%) |
| IDH-mutant, non-codeleted | 6 (29%) |
| IDH-wildtype | 10 (48%) |
| *Unknown* | *2 (10%)* |
| **Epilepsy present, n (%)** | 14 (67%) |
| *Unknown* | *3 (15%)* |
| **Preoperative tumor volume (cm$^3$), mean ± SD** | 49.1 ± 44.4 |
| **Days between craniotomy and MEG, median (Q1-Q3)** | 104 (95-463) |
| **Radiological follow-up around MEG** | |
| Days between baseline MRI and subsequent MEG, median (Q1-Q3) | 55 (18-166) |
| Days between MEG and subsequent MRI, median (Q1-Q3) | 92 (57-170) |
| **Treatment during MEG** | |
| Radiotherapy | - |
| Chemotherapy | 4 (19%) |
| Both | 2 (10%) |
| **Change in tumor volumes** | |
| Increasing tumor volume | 7 (33%) |
| Stable tumor volume | 14 (67%) |

* No MRIs available after MEG. SD = standard deviation, Q1-Q3 = 1st quartile-3rd quartile.

**Supplementary table 9: Correlations between tumor volume changes and global brain activity in the validation cohort**

| | *Correlations with change in tumor volume (ml)* | |
|---|---|---|
| | **All (n=21)** | **Increasing volume (n=7)** |
| **Global broadband power** | *Rho=0.07, P=0.763* | *Rho=0.25, P=0.589* |
| **Global offset** | *Rho=0.06, P=0.795* | *Rho=0.29, P=0.535* |
| **Global slope** | *Rho=0.09, P=0.713* | *Rho=0.43, P=0.337* |
| | *Correlations with change in tumor volume (%)* | |
| | **All (n=17)** | **Increasing volume (n=4)** |
| **Global broadband power** | *Rho=0.11, P=0.675* | *Rho=-0.80, P=0.200* |
| **Global offset** | *Rho=0.09, P=0.742* | *Rho=-0.80, P=0.200* |
| **Global slope** | *Rho=0.06, P=0.814* | *Rho=-0.80, P=0.200* |

*Note. Significance was determined after correction for the six tests performed per change score (ml or percentage).*

**Supplementary table 10: Global brain activity across groups in the validation cohort**

| | **Increasing volume, n=7** | **Stable volume, n=14** | **U-statistic, p-value** |
|---|---|---|---|
| **Global broadband power** | 3719 (3568-4009) | 3894 (3505-4391) | U=38, P=0.217 |
| **Global offset** | 5.79 (5.69-5.91) | 5.84 (5.72-5.97) | U=41, P=0.288 |
| **Global slope** | 0.94 (0.86-1.04) | 0.97 (0.91-1.00) | U=45, P=0.397 |

Note. Median (1st quartile-3rd quartile) is reported. Significance was determined after correction for the three tests performed.

**Supplementary figure 6: Brain activity of peritumoral and homologue contralateral areas in validation cohort**

*Peritumoral brain activity was significantly higher compared to the homologue contralateral areas for broadband power (W=28, P=0.001, D=0.56), offset (W=22, P=0.001, D=0.59) and slope (W=12, P<0.001, D=0.88). * P=0.001, ** P <0.001, significant after correction for the three tests performed. [A.U.] = arbitrary units.*

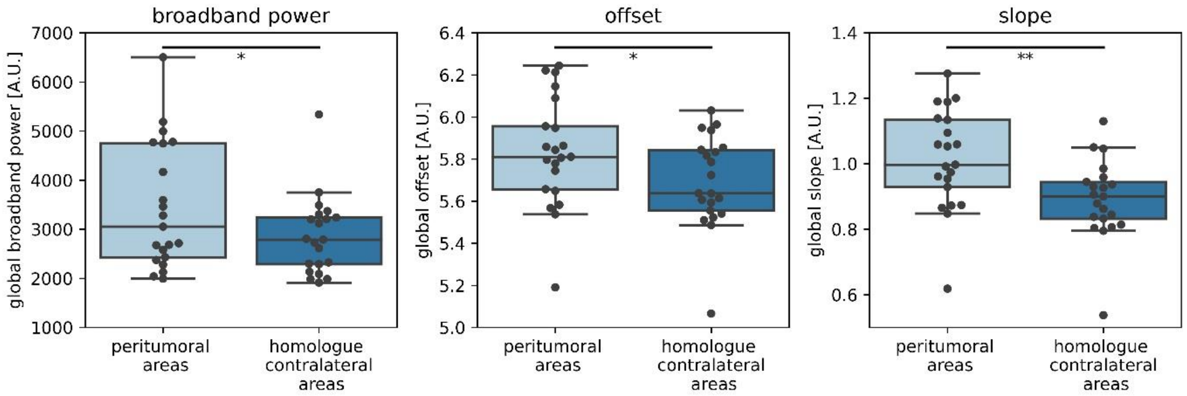

**Supplementary figure 7: Normalized peritumoral brain activity in the validation cohort**
*No significant differences were found between patients with increasing tumor volume or stable volume for broadband power ratio (U=40, P=0.263), offset ratio (U=45, P=0.397) or slope (U=31, P=0.096), after correction for the three tests performed. [A.U.] = arbitrary units.*

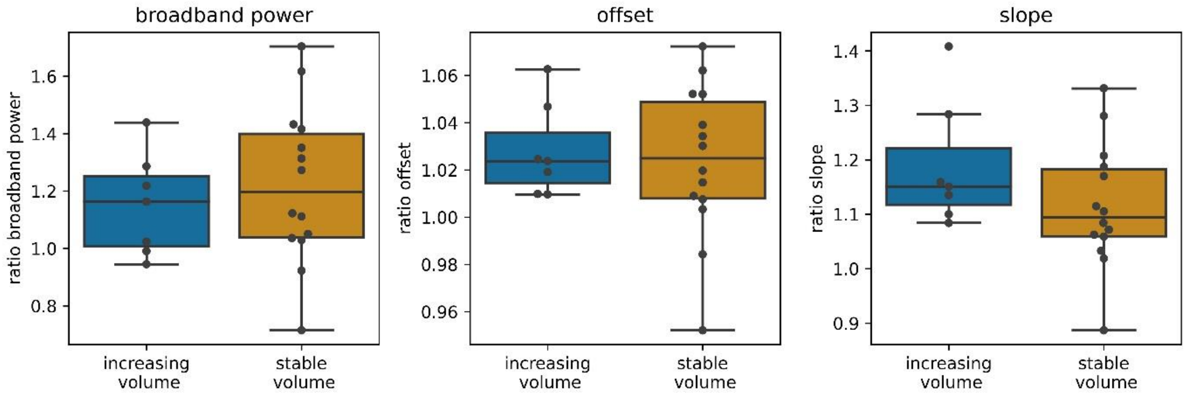


### Supplementary information MEEG cohort

#### *Methods*

Glioma patients of 18 years and older, without history of psychiatric or neurological diseases, who underwent craniotomy at the Amsterdam University Medical Centers (Amsterdam UMC, location VUmc) between June 2019 and February 2020 were eligible to participate. The World Health Organization 2016 classification of glioma was performed using histopathology and molecular classification.

MEG and EEG were recorded simultaneously. The same 306-channel whole-head Elekta system (Elekta Neuromag Oy, Helsinki, Finland) as described for the main cohort was used. The EEG recording was performed with a 60-channel MEG-compatible EEG cap, (Easycap Gmbh, Herrsching, Germany), with a sample frequency of 1250Hz, and hardware filters at 410Hz low-pass and 0.1Hz high-pass. See manuscript for MEG preprocessing and data selection.

Brain activity was computed at the sensor-level for the included EEG channels and at source-level for the MEG recordings, since we were mainly interested in sensor-level EEG as a more widely available and accessible technique to measure brain activity. Results were averaged across the ten selected epochs for each modality. Differences in brain activity of the hemisphere ipsilateral to the tumor with the contralateral hemisphere were assessed using Wilcoxon signed-rank tests, for EEG and MEG data separately.

*Results*

Seventeen patients were postoperatively included. One patient had to be excluded from final analysis because the EEG recording contained major artifacts. The majority of patients were grade IV glioblastoma (supplementary table 10).

The hemisphere ipsilateral to the tumor showed significantly higher brain activity compared to the hemisphere contralateral to the tumor for both MEG and EEG (supplementary figure 8).

**Supplementary table 11: Patient characteristics of MEEG cohort**

| | **MEEG cohort (n=16)** |
|---|---|
| **Age, mean ± SD** | 56.3 ± 11.1 |
| **Male, n (%)** | 11 (69%) |
| **WHO Grade, n (%)** | |
| II | 4 (25%) |
| IV | 12 (75%) |
| **Molecular subtype, n (%)** | |
| IDH-mutant, 1p/19q codeleted | 3 (19%) |
| IDH-mutant, 1p/19q status unknown | 1 (6%) |
| IDH-wildtype | 12 (75%) |
| **Epilepsy present, n (%)** | 11 (69%) |
| **Days between craniotomy and MEG, median (Q1-Q3)** | 56 (44-80) |
| **Lateralization of tumor, n (%)** | |
| Right | 8 (50%) |
| Left | 8 (50%) |

SD = standard deviation, Q1-Q3 = 1$^{st}$ quartile-3$^{rd}$ quartile

**Supplementary figure 8: Hemispheric brain activity of hemispheres in the MEEG cohort**
*Significantly higher brain activity was found in the ipsilateral hemisphere compared to the contralateral hemisphere to the tumor, with both EEG (broadband power: W=1, P<0.01, D=0.44; offset: W=12, P<0.01, D=0.52; slope: W=23, P=0.02, D=0.25) and MEG (broadband power: W=15, P<0.01, D=0.50; offset: W=10, P<0.01, D=0.84; slope: W=12, P<0.01, D=0.82). *P<0.05, ** P<0.01 after correction for the six the six comparisons drawn.*

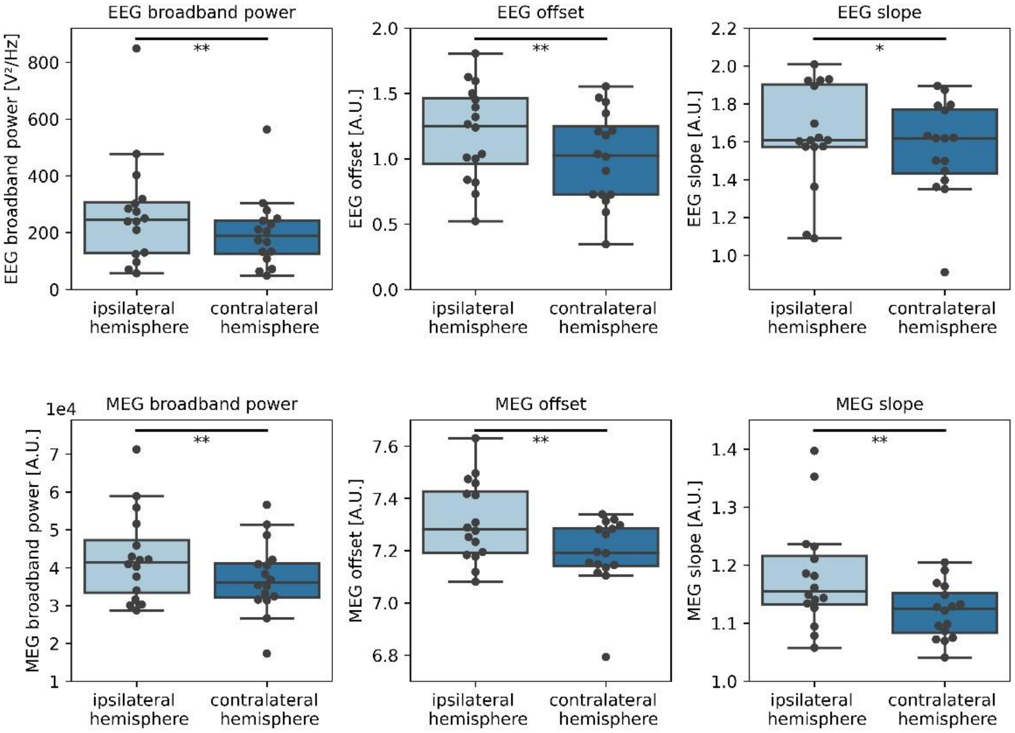


### Supplementary information assessing the robustness of epoch selection

To assess the effect of epoch selection on our results, the between-group comparison was repeated with ten epochs selected based on the highest alpha peak (maximum power in the 4-13 Hz band). Pearson's correlations of broadband power calculated using the two different selection methods was 0.96 (P<0.01). There were no differences in brain activity in stable versus increasing tumor volumes using the alpha peak-selected epochs (increasing tumor volume: median 3.42e+08 (IQR 3.11e+08-3.86e+01), stable tumor volume: median 3.57e+08 (3.32e+08-3.86e+08), U=223, P=0.33).

### Reference


1. Robinson S.E., Vrba, J. Functional neuroimaging by synthetic aperture magnetometry (SAM) (1998). In: Yoshimoto T, Kotani M, Kuriki S, Karibe H, Nakasato N, editors. Recent Advances in Biomagnetism. Sendai: Tohoku University Press.
